# Trends in COVID-19 vaccination intent, determinants and reasons for vaccine hesitancy: results from repeated cross-sectional surveys in the adult general population of Greece during November 2020-June 2021

**DOI:** 10.1101/2021.06.23.21259376

**Authors:** V Sypsa, S Roussos, V Engeli, D Paraskevis, S Tsiodras, A Hatzakis

**Author notes:** **Corresponding author** Vana Sypsa, Associate Professor of Epidemiology and Medical Statistics, Dept. of Hygiene, Epidemiology and Medical Statistics, Medical School, National and Kapodistrian University of Athens, Athens, Greece.

## Abstract

**Background:** Vaccine hesitancy is a major barrier to achieve large-scale COVID-19 vaccination. We report trends in vaccination intention and associated determinants from surveys in the adult general population in Greece.

**Methods:** Four cross-sectional phone surveys were conducted in November 2020, February, April and May 2021 on nationally representative samples of adults in Greece. Multinomial logistic regression was used on the combined data of the surveys to evaluate independent predictors of vaccination unwillingness/uncertainty.

**Results:** Vaccination intention increased from 67.6% in November 2020 to 84.8%. in May 2021. Individuals aged 65 years or older were more willing to get vaccinated (May 2021: 92.9% vs. 79.5% in 18-39 years, p<0.001) but between age-groups differences decreased over time. Vaccination intention increased substantially in both sexes, though earlier among men than women and was higher in individuals with postgraduate studies (May 2021: 91.3% vs. 84.0% up to junior high). From multivariable analysis, unwillingness and/or uncertainty to get vaccinated was associated with younger age, female gender (in particular in the April 2021 survey), lower educational level and living with a child ≤12 years old. Among those with vaccine hesitancy, concerns about vaccine effectiveness declined over time (21.6% in November 2020 vs. 9.6% in May 2021, p=0.014) and were reported more often by men; safety concerns remained stable over time (66.3% in November 2020 vs. 62.1% in May 2021, p=0.658) and were reported more often by women.

**Conclusions:** Vaccination intention increased substantially over time. Tailored communication is needed to address vaccine hesitancy and concerns regarding vaccine safety.

**Funding:** The phone surveys were conducted with the kind support of the Greek Shipowners’ Social Welfare Company SYN-ENOSIS

**Conflicts of interest statement:** The authors have no conflict of interest related to this article

## Introduction

Severe acute respiratory syndrome coronavirus 2 (SARS-CoV-2) has spread worldwide causing more than 172 million cases and 3.7 million deaths by early June 2021 [1]. Non pharmaceutical interventions have been employed worldwide to reduce COVID-19 healthcare demand and mortality. These interventions constitute a short term solution as their implementation over long periods has a major economic and social impact; in addition, the hardship associated with these measures resulted in “pandemic fatigue”, i.e. in gradual demotivation of people to follow recommended protective behaviours [2]. On the other hand, the continuing transmission of SARS-CoV-2 gives rise to new variants that may be more transmissible or more resistant to existing immunity and/or more pathogenic. As a result, developing safe and effective vaccines and achieving large-scale immunization is critical. Several vaccines have been approved by national regulatory authorities and more are under review [3]. High vaccination rates will be necessary to reach COVID-19 herd immunity, in particular with the emergence of more transmissible variants [4]. Apart from supply, administration and demand constraints that stand in the way in achieving high vaccination coverage rapidly [5], vaccine hesitancy is an important additional problem to be addressed.

The SAGE Working Group on Vaccine Hesitancy defined vaccine hesitancy as “the delay in acceptance or refusal of vaccination despite availability of vaccination services” [6]. The “3 Cs”, and more recently, the “5 Cs” models highlight concepts influencing the decision to get vaccinated such as Complacency, Confidence and Convenience/Constraints [6, 7]. Confidence, in particular, relates the trust in the effectiveness and safety of vaccines, the system that delivers them and the motivations of policy-makers who decide on the needed vaccines [6, 8]. Given the high levels of vaccination that are needed to achieve herd immunity, vaccine hesitancy poses a significant risk to minimizing the effects of the COVID-19 pandemic. Assessing the trends of vaccination intention over time and exploring its determinants could be of help to optimise communication and increase willingness to get vaccinated.

Our aim was to examine the trends in vaccination intention over time, and analyse determinants associated with vaccination hesitancy based on data from four cross-sectional surveys on nationally representative samples of adults in Greece. The surveys were implemented during the pandemic, from a period where vaccines were not available yet (November 2020) until approximately 5.5 million doses were distributed in the country population (May 2021).

## Methods

### Phone surveys and questionnaire

We conducted four cross-sectional phone surveys in the following periods: 17 November-3 December 2020, 1 February-18 February 2021, 1 April-12 April 2021 and 17 May-5 June 2021. In each survey, we recruited approximately 1,200 persons of all ages using proportional quota sampling based on age and sex from urban, semi-urban and rural areas in Greece. Questions concerning vaccination intention were asked only to adult respondents. Trained staff administered the questionnaires by phone.

The questionnaire included sections on sociodemographic characteristics (age, gender, education, occupation, nationality, area, household size, age of household members), social contacts, previous SARS-CoV-2 testing, vaccination intention and reasons for not intending to get vaccinated. Respondents were asked whether they would consider getting vaccinated - when it is their turn according to the national vaccination plan - with the following response categories: “Definitely yes”, “Probably yes”, “Probably not”, “Definitely not”, “Don’t know”, “Already vaccinated”. Those who responded “Probably not”/ “Definitely not” were asked to cite one or more reasons for not being willing to get vaccinated.

Vaccination roll-out in Greece is organised according to age. Other groups, such as healthcare workers and persons with underlying medical conditions, are also prioritised. In the periods when the surveys took place, vaccine availability was as follows: November 2020 – no vaccine; February 2021 – health care workers and persons aged 80 years or older; first half of April - individuals aged 60 years or older; second half of May - persons aged 30 years or older as well as 16-29 years old with underlying medical conditions (on May 29^th^, vaccination was available to all adults).

### Statistical methods

We estimated the proportions in each category of vaccination intention and the corresponding 95% confidence intervals (95% CI) after standardising for the age and sex distribution of the adult population in Greece.

We further assessed vaccination intention (“probably/definitely yes or already vaccinated”, “probably/definitely not”, “don’t know”) according to respondents’ characteristics and performed comparisons using the Chi-squared test.

A multinomial logistic regression model was used on the combined data of the four surveys to evaluate independent predictors of vaccination unwillingness and uncertainty. The outcome variable in this model was coded such that those likely to vaccinate (“probably/definitely yes or already vaccinated”) were compared to those who were i) unlikely to vaccinate (“probably/definitely not”) and ii) undecided about whether to vaccinate (“Don’t know”). Regression coefficients were exponentiated and are presented as relative risk ratios (RRR) with corresponding 95% confidence intervals (95% CI). We included interaction effects to assess changes in the impact of explanatory variables over time as well as to evaluate whether the impact of the variables was moderated by other characteristics of the respondents.

### Ethical issues

The protocol of the surveys was approved by the Institutional Review Board of the Hellenic Scientific Committee for the Study of AIDS and STDs. Participants provided oral informed consent.

## Results

### Characteristics of the participants

In the four surveys, individuals 18-39, 40-64 and 65+ years old constituted 25.5%-30.0%, 38.1%-45.9% and 28.2%-31.9% of the sample, respectively (Table 1). Approximately 53%-59% were women and 46%-53% had completed university or postgraduate studies. The median household size in the surveys was 2-3 persons; approximately one third and one fifth of the respondents lived in the same household with a person ≥65 years and a child≤12 years old, respectively.

**Table 1.**
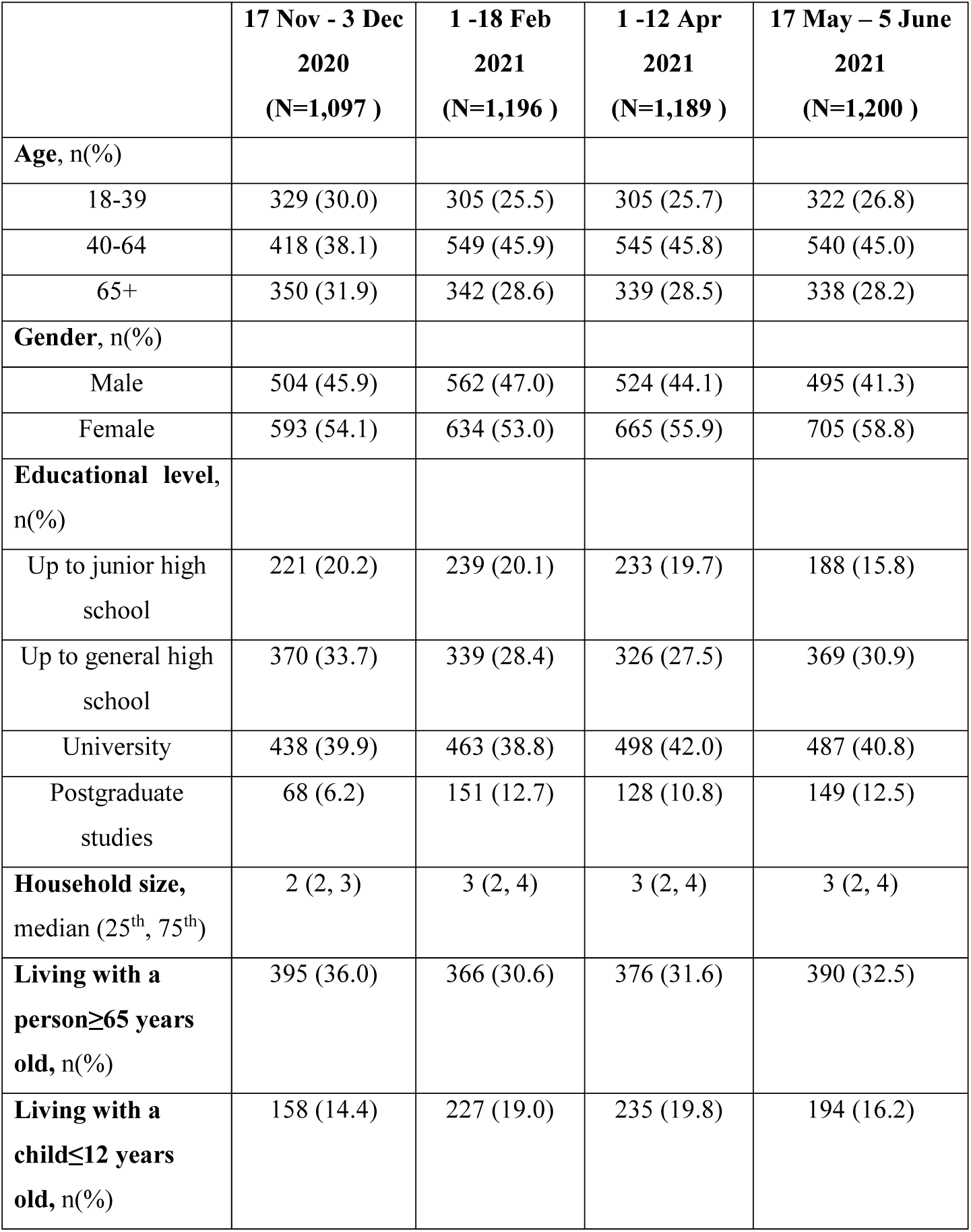
Socio-demographic characteristics of the participants in the 4 phone surveys in Greece

### Trends in vaccination intention

The trends in vaccination intention are depicted in Figure 1. There was a statistically significant increase over time in the proportion of those answering “Definitely yes” (including those already vaccinated) from 37.4% in November 2020 to 74.6% in May 2021. Across surveys, a decrease was observed in the proportion reporting “Probably yes” (from 30.2% to 10.2%) or “Don’t know” (from 13.2% to 2.5%) and a moderate reduction in those reporting “Probably not” (from 9.3% to 5.7%, respectively). Conversely, the proportion of participants answering “Definitely not” remained relatively stable over time (9.9% in November 2020 and 7.0% in May 2021).

**Figure 1.**
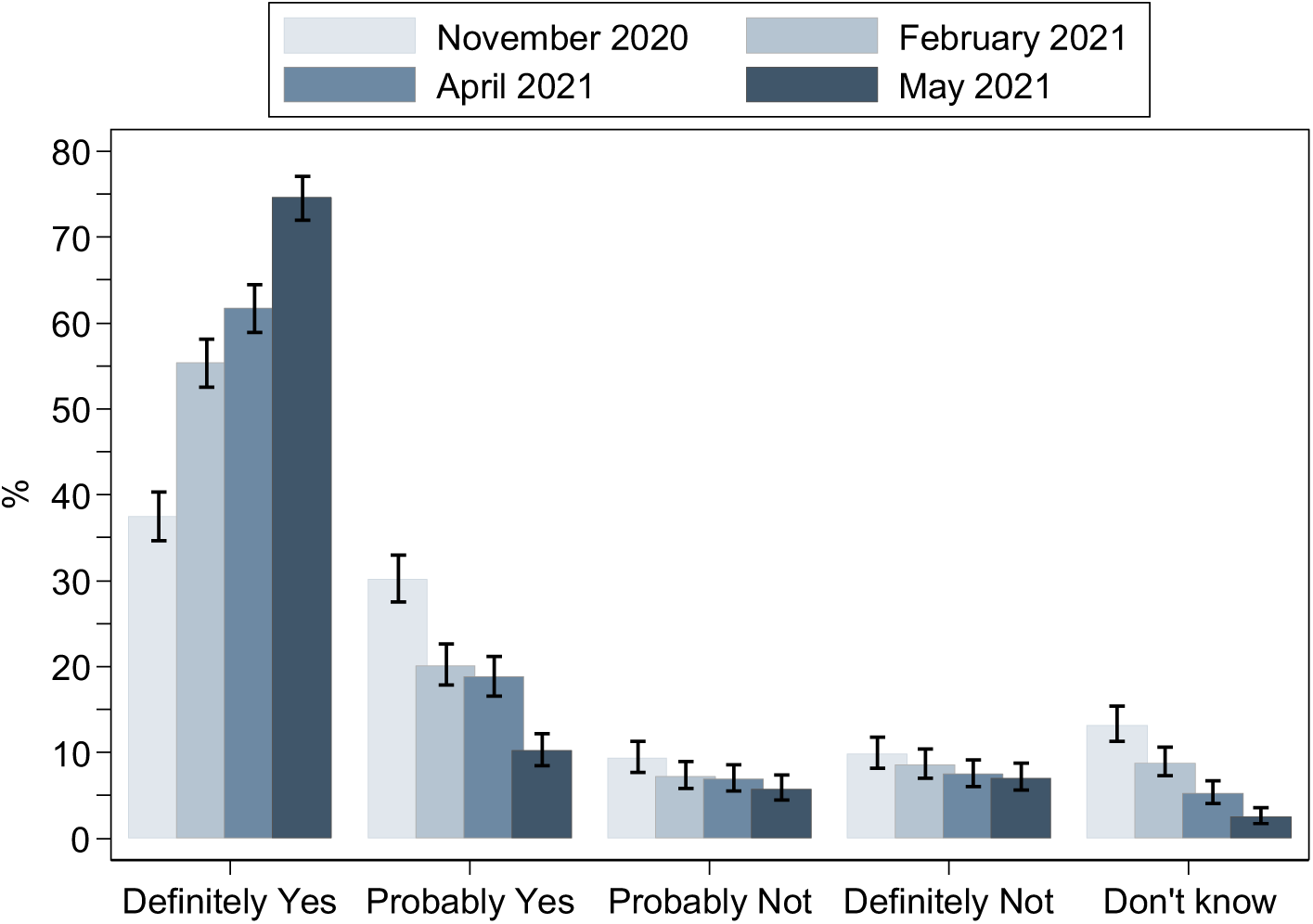
Responses concerning COVID-19 vaccination intention in four cross-sectional surveys implemented in the adult population of Greece during the pandemic (proportions and 95% Confidence Intervals). The category “Definitely yes” includes also those already vaccinated. The proportions are standardised to the age and sex distribution of the adult population in Greece.

### Trends in vaccination intention according to respondents’ characteristics

In November 2020, intent to receive COVID-19 vaccine (“definitely/probably yes”) was lower in younger individuals (58.7%, 68.9% and 79.1% in 18-39, 40-64 and 65+ years old, respectively, p<0.001) (Table 2). Over time, there was an increase in willingness to get vaccinated in all age groups and, although these differences were narrowed, they were still statistically significant in May 2021 (79.5%, 85.4% and 92.9% in 18-39, 40-64 and 65+ years old, respectively, p<0.001).

**Table 2.** Vaccination intention (% [95% Confidence interval]) over time by respondents’ characteristics (Willing: “Probably/Definitely yes” or “Already vaccinated”, Unwilling: “Probably/Definitely not”, Undecided: “Don’t know”)

|  | **17 Nov-3 Dec 2020**<br>**(N=1,097)** |  |  | **1 Feb -18 Feb 2021**<br>**(N=1,196 )** |  |  | **1 Apr-12 Apr 2021**<br>**(N=1,189 )** |  |  | **17 May-5 June 2021**<br>**(N=1,200 )** |  |  |
| --- | --- | --- | --- | --- | --- | --- | --- | --- | --- | --- | --- | --- |
|  | Willing | Unwilling | Undecided | Willing | Unwilling | Undecided | Willing | Unwilling | Undecided | Willing | Unwilling | Undecided |
| **Age group** |  |  |  |  |  |  |  |  |  |  |  |  |
| 18-39 | 58.7<br>[53.2, 63.9] | 26.7<br>[22.2, 31.8] | 14.6<br>[11.2, 18.8] | 66.9<br>[61.4, 72.0] | 23.6<br>[19.2, 28.7] | 9.5<br>[6.7, 13.4] | 75.7<br>[70.6, 80.2] | 18.0<br>[14.1, 22.8] | 6.2<br>[4.0, 9.6] | 79.5<br>[74.7, 83.6] | 18.6<br>[14.7, 23.3] | 1.9<br>[0.8, 4.1] |
| 40-64 | 68.9<br>[64.3, 73.2] | 17.9<br>[14.5, 21.9] | 13.2<br>[10.2, 16.8] | 76.3<br>[72.6, 79.7] | 14.4<br>[11.7, 17.6] | 9.3<br>[7.1, 12.0] | 79.6<br>[76.0, 82.8] | 15.0<br>[12.3, 18.3] | 5.3<br>[3.7, 7.6] | 85.4<br>[82.1, 88.1] | 11.1<br>[8.7, 14.1] | 3.5<br>[2.3, 5.5] |
| 65+ | 79.1<br>[74.6, 83.1] | 10.3<br>[7.5, 13.9] | 10.6<br>[7.8, 14.3] | 87.1<br>[83.1, 90.3] | 5.8<br>[3.8, 8.9] | 7.0<br>[4.7, 10.3] | 88.2<br>[84.3, 91.2] | 8.0<br>[5.5, 11.4] | 3.8<br>[2.2, 6.5] | 92.9<br>[89.6, 95.2] | 5.9<br>[3.8, 9.0] | 1.2<br>[0.4, 3.1] |
| **Gender** |  |  |  |  |  |  |  |  |  |  |  |  |
| Male | 69.8<br>[65.7, 73.7] | 18.1<br>[14.9, 21.7] | 12.1<br>[9.5, 15.3] | 78.5<br>[74.9, 81.7] | 13.3<br>[10.8, 16.4] | 8.2<br>[6.2, 10.8] | 84.4<br>[81.0, 87.2] | 12.4<br>[9.8, 15.5] | 3.2<br>[2.0, 5.2] | 85.7<br>[82.3, 88.5] | 11.1<br>[8.6, 14.2] | 3.2<br>[2.0, 5.2] |
| Female | 68.5<br>[64.6, 72.1] | 18.2<br>[15.3, 21.5] | 13.3<br>[10.8, 16.3] | 75.7<br>[72.2, 78.9] | 15.1<br>[12.6, 18.2] | 9.1<br>[7.1, 11.7] | 78.5<br>[75.2, 81.5] | 14.9<br>[12.4, 17.8] | 6.6<br>[5.0, 8.8] | 86.1<br>[83.3, 88.5] | 12.1<br>[9.8, 14.7] | 1.8<br>[1.1, 3.2] |
| **Educational level** |  |  |  |  |  |  |  |  |  |  |  |  |
| Up to junior high school | 66.5<br>[60.0, 72.4] | 15.4<br>[11.2, 20.8] | 18.1<br>[13.5, 23.8] | 71.1<br>[65.0, 76.5] | 15.5<br>[11.4, 20.7] | 13.4<br>[9.6, 18.3] | 78.1<br>[72.3, 83.0] | 13.7<br>[9.9, 18.8] | 8.2<br>[5.3, 12.4] | 84.0<br>[78.1, 88.6] | 12.8<br>[8.7, 18.4] | 3.2<br>[1.4, 6.9] |
| Up to high school | 70.8<br>[66.0, 75.2] | 15.9<br>[12.6, 20.0] | 13.2<br>[10.1, 17.1] | 72.6<br>[67.6, 77.1] | 17.1<br>[13.5, 21.5] | 10.3<br>[7.5, 14.1] | 77.9<br>[73.1, 82.1] | 16.3<br>[12.6, 20.7] | 5.8<br>[3.7, 9.0] | 83.5<br>[79.3, 86.9] | 13.8<br>[10.7, 17.7] | 2.7<br>[1.5, 5.0] |
| University | 68.5<br>[64.0, 72.7] | 20.8<br>[17.2, 24.8] | 10.7<br>[8.2, 14.0] | 81.0<br>[77.2, 84.3] | 13.4<br>[10.6, 16.8] | 5.6<br>[3.8, 8.1] | 81.9<br>[78.3, 85.1] | 13.9<br>[11.1, 17.2] | 4.2<br>[2.8, 6.4] | 87.3<br>[84.0, 90.0] | 11.1<br>[8.6, 14.2] | 1.6<br>[0.8, 3.3] |
| Postgraduate studies | 72.1<br>[60.2, 81.5] | 22.1<br>[13.7, 33.6] | 5.9<br>[2.2, 14.8] | 84.1<br>[77.3, 89.1] | 9.3<br>[5.6, 15.1] | 6.6<br>[3.6, 11.9] | 91.4<br>[85.1, 95.2] | 7.8<br>[4.2, 14.0] | 0.8<br>[0.1, 5.4] | 91.3<br>[85.5, 94.9] | 6.7<br>[3.6, 12.1] | 2.0<br>[0.6, 6.1] |

In November 2020, similar percentages in vaccination intent were identified between men and women (69.8% and 68.5%, respectively, p=0.821) and across educational levels (up to junior high: 66.5%, high school: 70.8%, university: 68.5%, postgraduate studies: 72.1%, p=0.670). Over time, willingness to get vaccinated increased substantially in both sexes, though earlier among men than women, and vaccination intention was similar in the most recent survey of May 2021 (85.7% of men and 86.1% of women) (Table 2). Vaccination intention increased substantially in all educational levels but this increase was more pronounced in university graduates or with postgraduate studies (May 2021: 84.0%, 83.5%, 87.3% and 91.3% for up to junior high, high school, university and postgraduate, respectively, p=0.036).

In all surveys, respondents who lived in the same household with a person 65 years or older were more likely to report willingness to get vaccinated as compared to those living alone or with younger individuals. This difference was narrowed in the most recent survey but remained statistically significant (May 2021: 88.7% vs. 84.6%, respectively, p=0.053). In the first survey in particular, where individuals 18-39 years old had substantially lower vaccination intention compared to other groups, young individuals living with an older person were more probable to be willing or indecisive as compared to those not living with an older person (83.3% vs. 70.1%, p=0.021).

Living with a child ≤12 years old was associated with higher vaccination refusal in the first surveys (November: 26.6% vs. 16.7%, February: 21.2% vs. 12.7%,) but there was no difference identified in the most recent surveys in April and May 2021. A closer look to the data revealed that this finding was evident among female respondents. In November 2020 for example, vaccination refusal for those living with a child vs. those without was 21.3% vs. 17.5%, for men (p=0.180) and 31.1 vs. 16.1% (p<0.001) for women.

### Predictors of unwillingness to get vaccinated or uncertainty with regards to vaccination

The results from the multinomial logistic regression model for the determinants of unwillingness or uncertainty about getting vaccinated are shown in Figure 2. The period when the survey took place had an effect with decreased vaccine hesitancy over time; compared to May 2021, respondents in November 2020 were more likely to be unwilling or uncertain and respondents in February 2021 were more likely to be uncertain. Groups at increased risk for unwillingness and uncertainty to vaccinate against COVID-19 were younger individuals (18-39 vs. 65+: unwilling: RRR = 7.1; 95% CI: 4.6 to 11.1, uncertain: RRR = 4.0; 95% CI: 2. 4 to 6.6 and 40-64 vs. 65+: unwilling: RRR = 4.3; 95% CI: 2.8 to 6.7, uncertain: RRR = 2.5; 95% CI: 1.5 to 4.1) as well as individuals with lower educational level (up to junior high vs. postgraduate: unwilling: RRR = 2.7; 95% CI: 1.9 to 3.9, uncertain: RRR = 4.3; 95% CI: 2.5 to 7.4, High school vs. postgraduate: unwilling: RRR = 2.1; 95% CI: 1.5 to 3.0, uncertain: RRR = 2.6; 95% CI: 1.5 to 4.3, University vs. postgraduate: unwilling: RRR = 1.8; 95% CI: 1.3 to 2.5, uncertain: RRR = 1.6; 95% CI: 0.9 to 2.7). Women were more likely to be unwilling as compared to men; this finding was more apparent among women aged 65 years or older (Figure 3A). Living with a child≤12 years old was unrelated to uncertainty around the COVID-19 vaccine but was associated with unwillingness (RRR vs. no child at home: 1.3; 95% CI: 1.1 to 1.6). There was an interaction between gender and survey period with women experiencing lower willingness and higher uncertainty in April 2021 as compared to men (Figure 3B).

**Figure 2.**
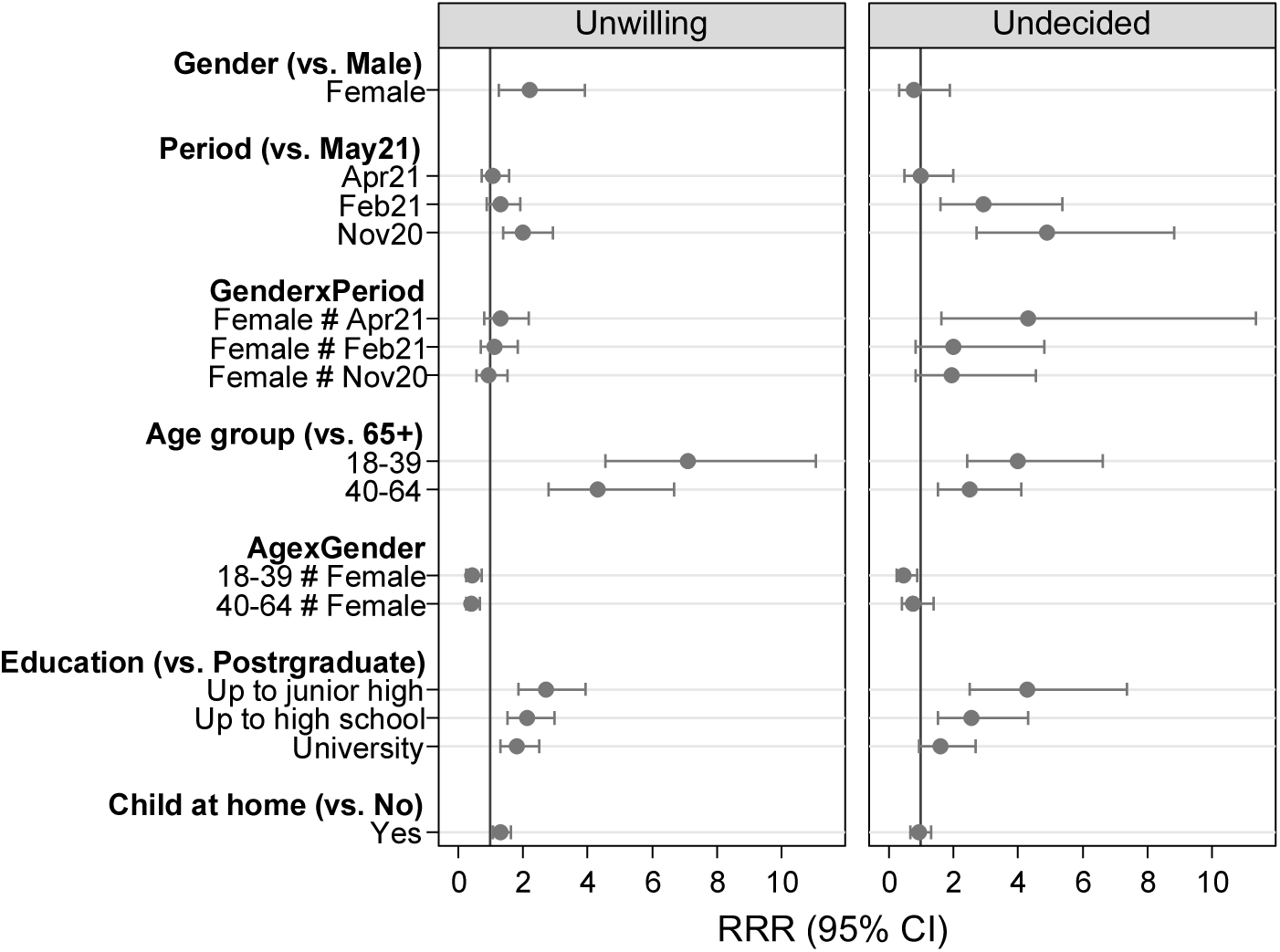
Predictors of unwillingness and uncertainty to vaccinate against COVID-19 based on the data collected from the four cross-sectional surveys in Greece (results from multivariable analysis)

**Figure 3.**
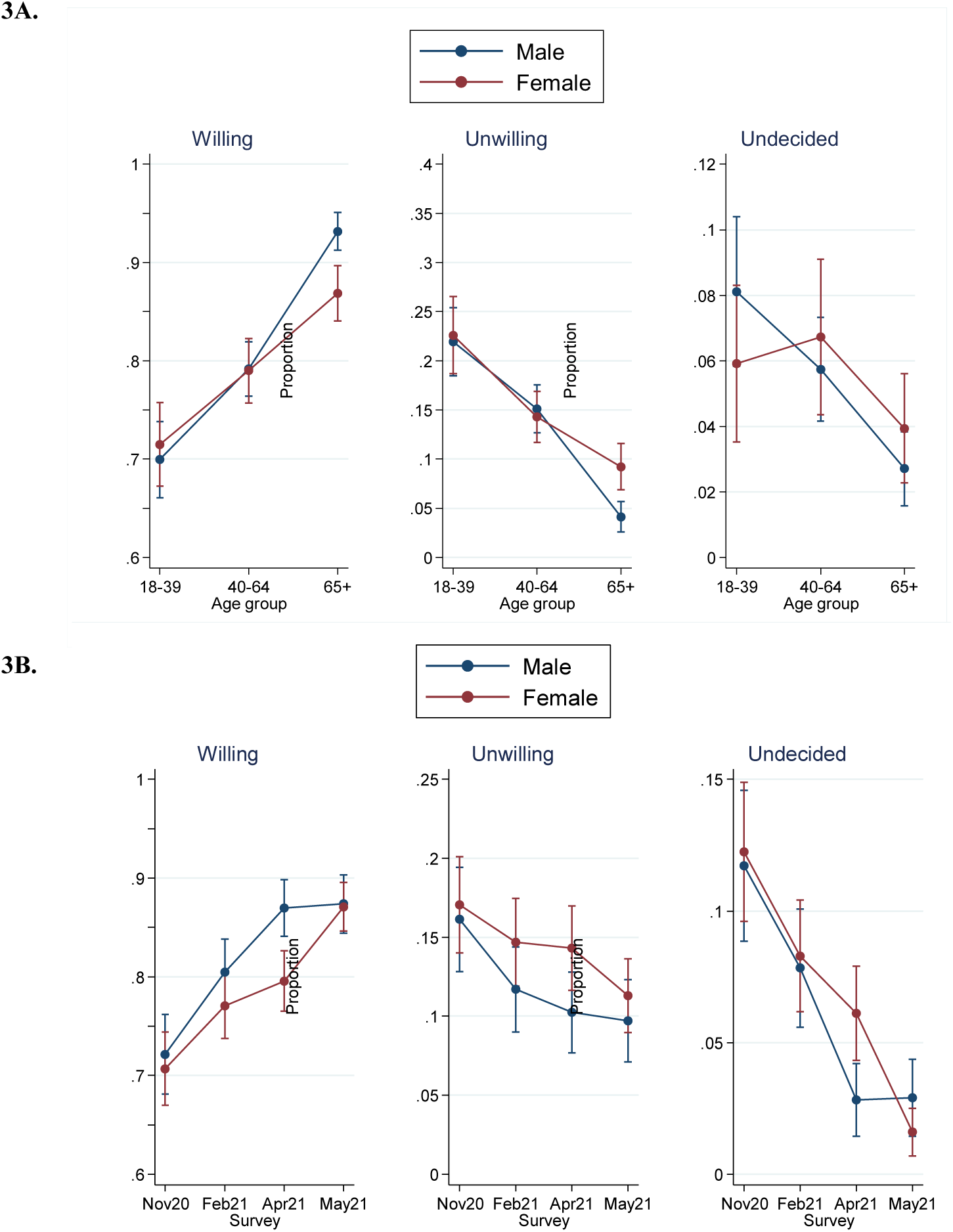
Adjusted predictions of willingness, unwillingness and uncertainty about getting vaccinated: **3A**. Effect of age according to the gender of respondents, **3B**. Effect of survey period according to the gender of respondents

### Reasons for not intending to get vaccinated

The reasons most frequently cited for not intending to get vaccinated were concerns about the safety and the effectiveness of the vaccine (65.5% and 15.7%, respectively). A larger percentage of the February, April and May surveys participants than November participants reported effectiveness concerns (21.6%, 17.1% and 15.0% vs. 9.6%, respectively, p=0.014). Safety concerns were reported at similar percentages in all four periods (66.3%, 62.6%, 67.7% and 62.1% in November 2020, February, April and May 2021, p=0.658). Some respondents specifically cited allergies as a concern relating to the safety of the vaccine in the beginning of vaccination process in the country but this concern diminished as vaccination progressed (fear of allergies among unwilling/uncertain: 0%, 7.6%, 2.4% and 0.7% in November 2020, February, April and May 2021, respectively, p<0.001).

Men more often reported doubts in vaccine effectiveness (19.9% vs. 12.4% reported by women, p=0.007) whereas women reported at higher percentages concerns about safety as compared to men (67.8% vs. 60.8%, p=0.062). The latter finding was modified by the age of the respondents; doubts about safety were reported more often by women aged 40-64 years old as compared men and this finding was independent of the educational level and the presence of a child in the household (Supplementary Figure 1).

As to other reasons for not intending to get vaccinated, 8.6% reported that they considered themselves as not being at risk of getting COVID-19 and 5.0% that COVID-19 is a mild disease.

## Discussion

We provide estimates on vaccination intention from four surveys conducted in the general adult population in Greece before the initiation of vaccination as well as during vaccination roll-out. A literature review on vaccination intention by October 2020 has found a substantial variation in vaccination intention between countries before vaccination was available [9]. Our estimate of 67.6% in November 2020 suggests that vaccination intention in the general adult population in Greece was high. It compares to a range of estimates between 59%-70% in US, Australia, Finland and UK from surveys performed between July-December 2020 [10-14] and lies between the estimates of 71.5% and 54% reported by surveys performed in 19 and 15 countries in June 2020 and January 2021, respectively [15, 16]. A survey performed in September-October 2020 in the personnel of healthcare facilities in Greece reported lower vaccination intention (51.1%)[17]; this however reflects attitudes in a sample with different age and sex distribution than that of the general adult population.

Vaccination intention in the last survey conducted in May 2021 increased to 84.8%. By that time, persons aged 30 years or older as well as 16-29 years old with underlying medical conditions had access to vaccination and, at the end of May, vaccination was offered to all adults. On May 31^st^, 2021, approximately 19% of the Greek population (23% of adults) was fully vaccinated. The most striking change from November 2020 to May 2021 was the 2-fold increase in those who were certain to receive vaccination. This increase seems to result from a shift of those who were initially positive towards vaccination (“Probably yes”) or undecided, as these proportions declined substantially over time; interestingly, the proportion of participants answering “Definitely not” remained relatively stable over time and at low numbers. Although willingness to get vaccinated among adults in May 2021 was high, this does not ensure that the high vaccination rates needed for herd immunity can be reached. First of all, vaccination intention is likely to be greater than actual vaccine uptake due to an “intention-behaviour gap” [18]. Furthermore, the vaccination of children remains controversial, in particular due to ethical dilemmas as health workers and vulnerable adults in other countries have not been yet vaccinated [19]. Despite that, the high vaccination intention observed in our survey among individuals 65 years and older (92.9%) is promising concerning achieving the target of reducing morbidity, mortality and healthcare burden. It remains to be seen whether such high rates of vaccination in this age group can be achieved in real-life.

In November 2020, i.e. before the initiation of vaccination, willingness to get vaccinated did not differ according to educational level or gender; it was higher in older individuals and people living with elderly and lower in people living with a child. Subsequently, an increase in vaccination intention was apparent in all age groups and, in particular, among individuals 18-39 years old; as a result, differences according to age were narrowed over time but were not eliminated as people 65 years or older were still more willing to get vaccinate. Apart from younger age, women were more likely to be unwilling or uncertain about getting vaccinated and that was independent of educational level. This finding has been reported in the literature [10, 11, 13, 20, 21]. We have identified effect modification of the relationship between age and vaccination intention by gender; women 65 years or older were less likely to accept vaccination compared to males. Also, women exhibited higher uncertainty during vaccination roll-out (in particular in the survey of April 2021) and that was independent of their educational level and their age. This was most probably caused by information on vaccine-linked blood clots being announced by the European Medicines Agency in early April [22]. The finding on the impact of gender deserves attention as, apart from the fact that women constitute half of the world population and account for a large proportion of healthcare workers [23], they also influence health care decisions in their families.

Higher unwillingness and uncertainty was identified also among respondents with lower educational level. Educational levels, as well as related social characteristics, such as income, have been found to affect attitudes towards COVID-19 vaccination also elsewhere [11, 12, 20, 24, 25]. In the US in particular, based on the data from the “Understanding Coronavirus in America” survey, it became evident that education level played a greater role in people’s willingness to get the vaccine than race and ethnicity [26]. This finding shows that people with lower educational level constitute a group that cannot be easily convinced by conventional risk communication and additional effort is needed. We have also found that living with a child≤12 years old was associated with higher probability of vaccine refusal. This has been reported in another study as well [11]. Probably, parents who are not convinced about the safety of the vaccine are more conservative in taking a potential risk.

Based on our findings, there is a core of people who are not convinced about the effectiveness and the safety of the vaccine and the size of this population – in particular those refusing vaccination - did not change substantially during the expansion of vaccination around the world. In our studies, approximately two-thirds of those not intending to get vaccinated reported as main reason concerns about the safety of the vaccine and 16% about its effectiveness. Although doubts about vaccine effectiveness decreased over time, safety concerns remained at similarly high levels. It is of note that there were gender differences with men reporting more often doubts in vaccine effectiveness and women reporting at higher percentages concerns about its safety. This explains why although men and women did not differ in their vaccination intention in November, over time men became more quickly convinced and were more willing to get vaccinated whereas women were more often negative or undecided.

Our study has some limitations. First, selection bias could exist in the phone surveys. Interviewers informed the persons who picked up the phone that they were going to ask questions about COVID-19. It is possible that people with negative attitudes towards the pandemic would be more probable to decline. Second, responses might be affected by social desirability bias; for example, respondents might answer questions about vaccination intention in a manner that would be viewed favourably by the interviewers. Both these biases might result in overestimating vaccination intention. However, the findings concerning trends in vaccination intention over time and associated determinants should not be affected by this limitation. Third, the findings on vaccination intention over time and associated determinants could be influenced by the disease burden in the country, social distancing measures or media coverage during the specific survey period. This is not necessarily a limitation as, for example, it allowed us to assess the impact of news on vaccine-linked blood clots on vaccination intention among women.

A strength of this study is that vaccination intention and its determinants have been evaluated through four repeated cross-sectional surveys implemented over a period extending from before vaccination was available until it was offered to all the adults in the country. As persons aged 18 years or older constitute approximately 83% of the population in Greece, a vaccination intention of 84% could result in 69% of the total population accepting the vaccine. To ensure that we reach this target, it is important to address convenience and constraints for those willing to get vaccinated by removing practical barriers to vaccination and improving ease of access in order to close the potential “intention-behaviour gap”. A major concept influencing the decision to get the COVID-19 vaccine is confidence; people who refuse or are uncertain about getting vaccinated usually express concerns about the safety of the vaccine. Tailored communication that takes into account the characteristics of the population with lower vaccine acceptance and providing consistent and accurate information are needed to address vaccine hesitancy and safeguard the public against vaccine misinformation.

## Data Availability

Data are not available for sharing.

## Authorship

VS and AH designed the surveys. VS, SR and VE reviewed the literature and analysed the data. VS, SR, VE, DP, ST and AH interpreted the data. VS wrote the first draft. SR, VE, DP, ST and AH revised the draft for important intellectual content

---

**Supplementary Figure 1.**
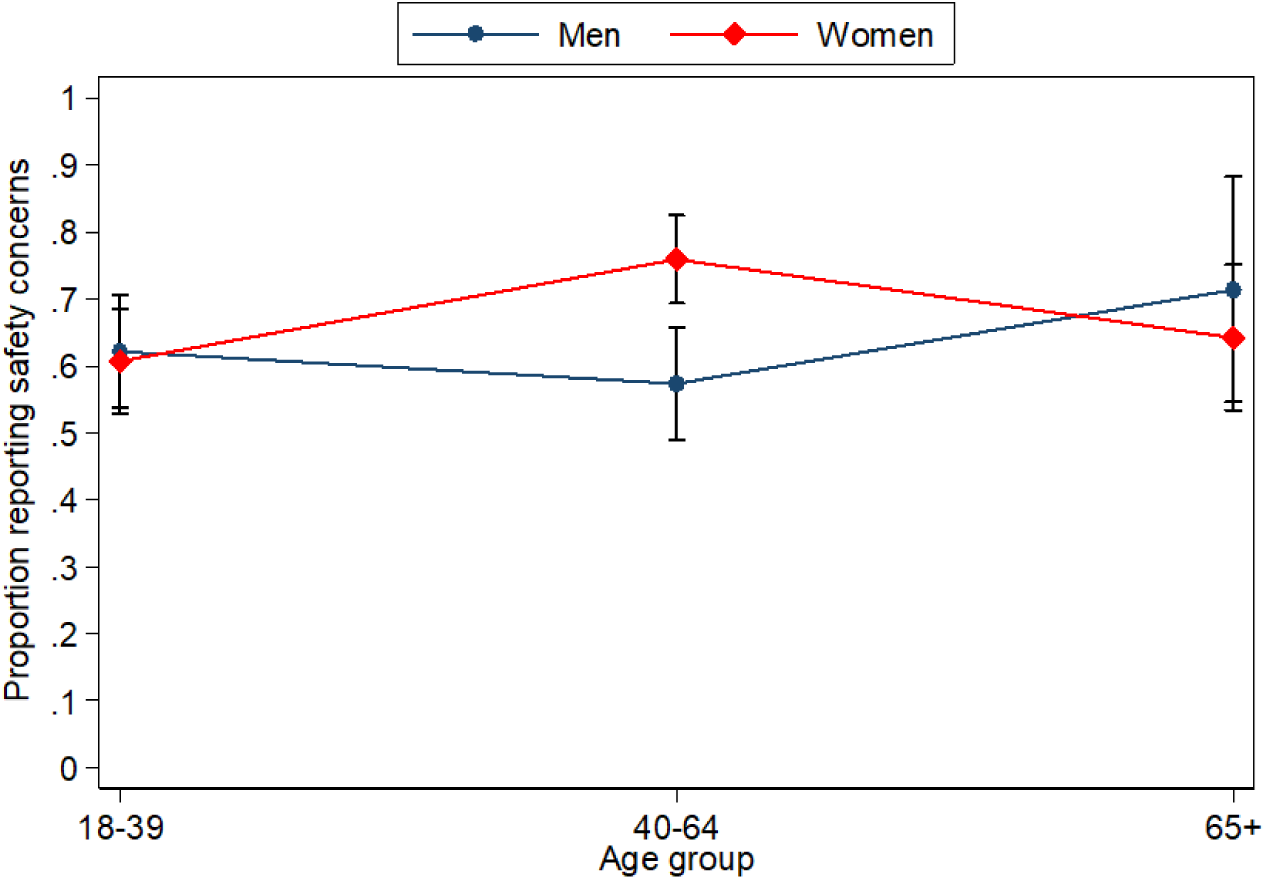
Adjusted predictions of reporting safety concerns by gender and age group (among respondents unwilling to get vaccinated)

